# Trends and Future Projections of Prevalence and YLDs Rate of Heart Failure in Asia: A Systematic Analysis Based on GBD 2023

**DOI:** 10.64898/2026.04.29.26352089

**Authors:** Guoping Gu, Ping Yang, Yong Li

**Author notes:** **Correspondence to:** Guoping Gu.

## Abstract

**Objective:** To analyze the burden, trends, health inequalities, and future projections of heart failure in Asia from 1990 to 2023 based on the Global Burden of Disease (GBD) 2023 study.

**Methods:** Age-standardized prevalence rates (ASPR) and age-standardized Years Lived with Disability (YLDs) rates were calculated. Joinpoint regression, Age-Period-Cohort (APC) models, Das Gupta decomposition analysis, Data Envelopment Analysis (DEA), Slope Index of Inequality (SII), Concentration Index (CI), and Bayesian Age-Period-Cohort (BAPC) models were employed.

**Results:** In 2023, approximately 30.29 million (95% uncertainty intervals [UI]: 23.66–37.99 million) heart failure cases occurred in Asia, with an ASPR of 602.603 per 100,000 (95% UI: 471.499–754.961). YLDs totaled 2.92 million (95% UI: 1.89–4.26 million), with an age-standardized YLDs rate of 57.970 per 100,000 (95% UI: 37.575–84.627). From 1990 to 2023, ASPR increased with an estimated annual percentage change (EAPC) of 0.252% (95% CI: 0.231–0.273) and the age-standardized YLDs rate increased with an EAPC of 0.238% (95% CI: 0.219–0.257). East Asia had the highest ASPR (674.809 per 100,000) and YLDs rate (65.802 per 100,000). Males had higher ASPR and YLDs rates than females across all subregions. Decomposition analysis showed that aging (49.04%) and population growth (41.80%) were the primary drivers of burden increase. SII deteriorated from 10.684 in 1990 to 25.003 in 2023 (134.0% increase), and CI declined from −0.077 to −0.229, indicating widening health inequalities. BAPC projections estimated that ASPR will rise to 623.351 per 100,000 and the age-standardized YLDs rate to 60.596 per 100,000 by 2038.

**Conclusions:** Heart failure burden in Asia increased from 1990 to 2023 with marked regional and gender disparities, expanding health inequalities and is projected to continue rising through 2038.

## 1 Introduction

Heart failure is a complex clinical syndrome that affects multiple organs and systems throughout the body, representing a leading cause of cardiovascular morbidity and mortality worldwide ^[1–3]^. The most common etiologies of heart failure include hypertension, coronary artery diseases, diabetes mellitus, obesity, smoking, and genetic factors ^[4].^ Currently, approximately 64 million individuals globally suffer from heart failure, imposing an enormous burden on healthcare systems with annual medical costs exceeding $346 billion ^[5, 6].^ With the intensification of population aging and the prevalence of cardiovascular risk factors, heart failure has emerged as a critical public health concern.

Although heart failure medications continue to be developed and treatment approaches continue to advance, the five-year survival rate following heart failure diagnosis remains below 50%, making heart failure one of the leading causes of death in many countries ^[7–9]^. The prevalence of heart failure in Western countries is estimated to range from 1% to 14%, reflecting significant geographic heterogeneity in the disease burden of heart failure ^[5, 10]^. With the relative contributions of factors such as population aging, the prevalence of risk factors including hypertension and diabetes, and improvements in diagnostic capabilities in Asian regions, the prevalence of heart failure continues to rise. However, dedicated research on the characteristics of heart failure disease burden in Asia remains relatively insufficient, with most existing studies focusing on descriptive analyses and lacking multidimensional comprehensive assessment approaches.

This study represents the first comprehensive analysis of heart failure disease burden in Asia based on the latest GBD 2023 data, with coverage extending to 2023. The study innovatively integrates multiple advanced epidemiological analytical methods and focuses specifically on the Asian region, fully accounting for the unique characteristics of Asia’s population structure, economic development, and cultural context. It elucidates the association between heart failure disease burden and socioeconomic development levels, examines the phenomenon of persistently worsening health inequalities and their underlying mechanisms, and provides a novel analytical perspective for prevention and control efficiency assessment, thereby offering crucial prospective evidence to support policy formulation.

## 2 Methods

### 2.1 Data Sources and Indicator Calculations

This study utilized heart failure epidemiological data for Asia from 1990 to 2023, obtained from the Global Burden of Disease (GBD) 2023 database via the GBD official website (https://vizhub.healthdata.org/gbd-results/). In accordance with the GBD 2023 regional classification framework, “Asia” in the present study includes specifically to the four GBD-defined sub-regions:East Asia, Southeast Asia, Central Asia, and South Asia. It should be noted that countries conventionally regarded as belonging to West Asia (the Middle East) are classified under the “Middle East and North Africa (MENA)” super-region within the GBD database, and are therefore not included in the present analysis. Corresponding demographic data were obtained for standardization calculations. The primary calculated indicators included age-standardized prevalence rates (ASPR) and age-standardized YLDs rates. The GBD 2023 study employs a standardized data collection and estimation framework to ensure internal consistency and comparability across countries and over time. Heart failure is defined according to ICD-10 codes (I50), applied consistently across all locations and study years. Input data are drawn from multiple sources, including vital registration systems, hospital discharge records, disease registries, household surveys, and published epidemiological literature, all of which undergo systematic quality assessment and bias adjustment prior to modeling. Disease burden estimates are generated using DisMod-MR 2.1, a Bayesian meta-regression tool that simultaneously models incidence, prevalence, remission, and mortality within a compartmental framework, thereby enforcing biological and epidemiological consistency across age groups, sexes, geographies, and calendar years. All estimates are reported with 95% uncertainty intervals (UI), which integrate both sampling uncertainty and model uncertainty. Countries or regions with sparser or lower-quality input data are assigned correspondingly wider UIs, providing an objective indicator of data reliability at the country level.

### 2.2 Descriptive analysis of disease burden

Descriptive analysis of heart failure incidence and YLDs rates in Asia from 1990-2023 was conducted, calculating prevalence cases, YLDs numbers, ASPR, and age-standardized YLDs rates for each region and year, as well as percentage changes in these indicators from 1990-2023. Disease burden heat maps were created to display heart failure distribution in Asia.

### 2.3 Joinpoint regression analysis

Joinpoint regression models were applied to analyze trends in heart failure age-standardized rates from 1990-2023. Standard errors of age-standardized rates were calculated using R software, data were imported into Joinpoint software with time as the independent variable, age-standardized rates as the dependent variable, and region as the grouping variable, selecting a maximum of 5 joinpoint points with 95% confidence intervals for parameter testing. Results were expressed as Annual Percent Change (APC).

### 2.4 Age-Period-Cohort analysis

To explore temporal trends in heart failure, data from 1990-1993 were excluded, and 1994-2023 data were divided into 5-year periods and uploaded to websites for age, period, and cohort effect analysis. Since the GBD database reports age-group data in 5-year intervals, the APC model correspondingly requires time periods to be divided into equal-length 5-year intervals to maintain methodological consistency. Therefore, data from 1990 to 1993 were excluded, as including these years would have produced an incomplete 4-year initial interval (1990–1993) that violates the equal-interval assumption fundamental to valid APC estimation. Effects of each factor on heart failure were calculated separately, and relative risk change graphs were plotted for age, period, and cohorts. Local drift percentages for disease risk in each age group compared to the previous age group were calculated.

### 2.5 Decomposition analysis

Das Gupta decomposition method was applied to analyze the effects of demographic factors, epidemiological factors, and population age structure factors on heart failure disease burden changes. Analysis was conducted separately for Asia, different genders, and different Asian regions.

### 2.6 Frontier analysis

To deeply explore the association between heart failure disease burden and social development levels, this study employed Data Envelopment Analysis (DEA) methods, selecting 32 Asian countries and regions with complete Human Development Index (HDI) data from 1990-2023 as research samples for frontier efficiency analysis of HDI and heart failure age-standardized YLDs rates. The study used Free Disposal Hull (FDH) models to construct nonlinear production frontiers and employed Locally Weighted Scatterplot Smoothing (LOWESS) to generate smooth frontier curves. To ensure robustness of analytical results, super-efficient observation points (outlier points below the frontier boundary) were excluded. The constructed DEA efficiency frontier reflected theoretically achievable optimal heart failure YLDs rates at given HDI levels, providing scientific efficiency benchmarks and comparison standards for objectively evaluating national heart failure prevention and control performance.

### 2.7 Health inequality analysis

To assess health equity in heart failure disease burden, this study employed Slope Index of Inequality (SII) and Concentration Index (CI) to analyze distribution characteristics of heart failure YLDs rates in 1990 and 2023. SII reflected absolute inequality in disease burden, while CI reflected relative inequality. SII and CI for 1990 and 2023 were calculated separately, scatter plots were created, and health inequality changes between the two time points were compared.

### 2.8 Heart failure prevalence prediction analysis

Bayesian Age-Period-Cohort (BAPC) models were used to project heart failure ASPR and age-standardized YLDs rates for the period 2024-2038. These projections were based on the extrapolation of historical trends and assumed that past age, period, and cohort patterns continue unchanged into the future. BAPC models considered age, period, and cohort effects, setting prior distributions and combining sample data for posterior inference of the three effects, finally combining the three effects to generate projected disease burden trajectories.

### 2.9 Statistical analysis

All data analysis and visualization were completed using R 4.2.0. Statistical significance was set at *P* < 0.05.

## 3 Results

### 3.1 Trends of Heart Failure Disease Burden in Asia

In 2023, approximately 30.29 million heart failure cases occurred in Asia (95% UI: 23.66-37.99 million), with an age-standardized prevalence rate (ASPR) of 602.603/100,000 (95% UI: 471.499-754.961). Among the four subregions in Asia, East Asia had the highest ASPR (674.809/100,000), followed by Central Asia (659.401/100,000), while South Asia had the lowest (493.527/100,000). At the country level, United Arab Emirates had the highest ASPR (931.240/100,000), while Brunei had the lowest (448.820/100,000). In 2023, heart failure-related Years Lived with Disability (YLDs) in Asia totaled approximately 2.92 million (95% UI: 1.89-4.26 million), with an age-standardized YLDs rate of 57.970/100,000 (95% UI: 37.575-84.627). East Asia had the highest age-standardized YLDs rate (65.802/100,000), while South Asia had the lowest (46.022/100,000). At the country level, United Arab Emirates had the highest age-standardized YLDs rate (87.773/100,000), and Brunei had the lowest (42.040/100,000) (Table 1).

**Table 1.**
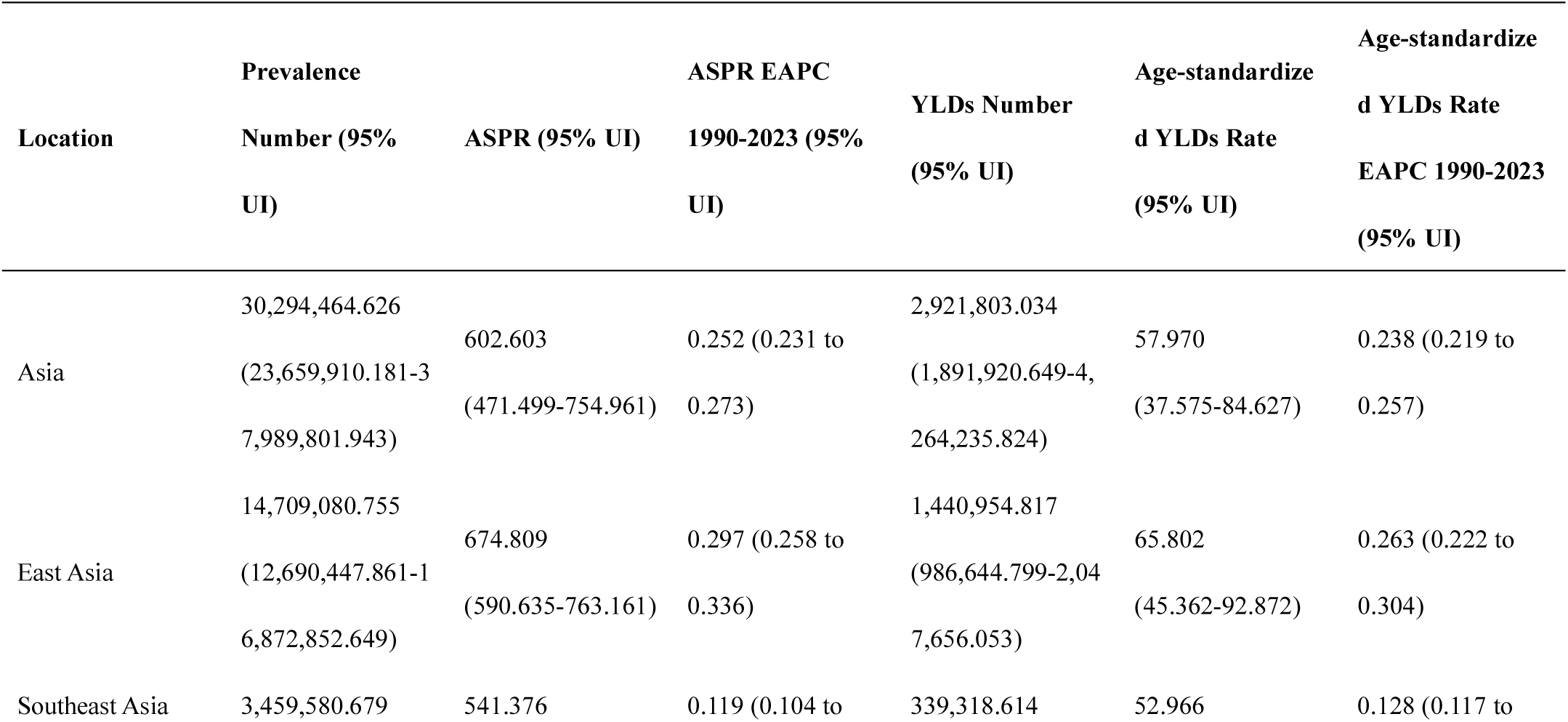
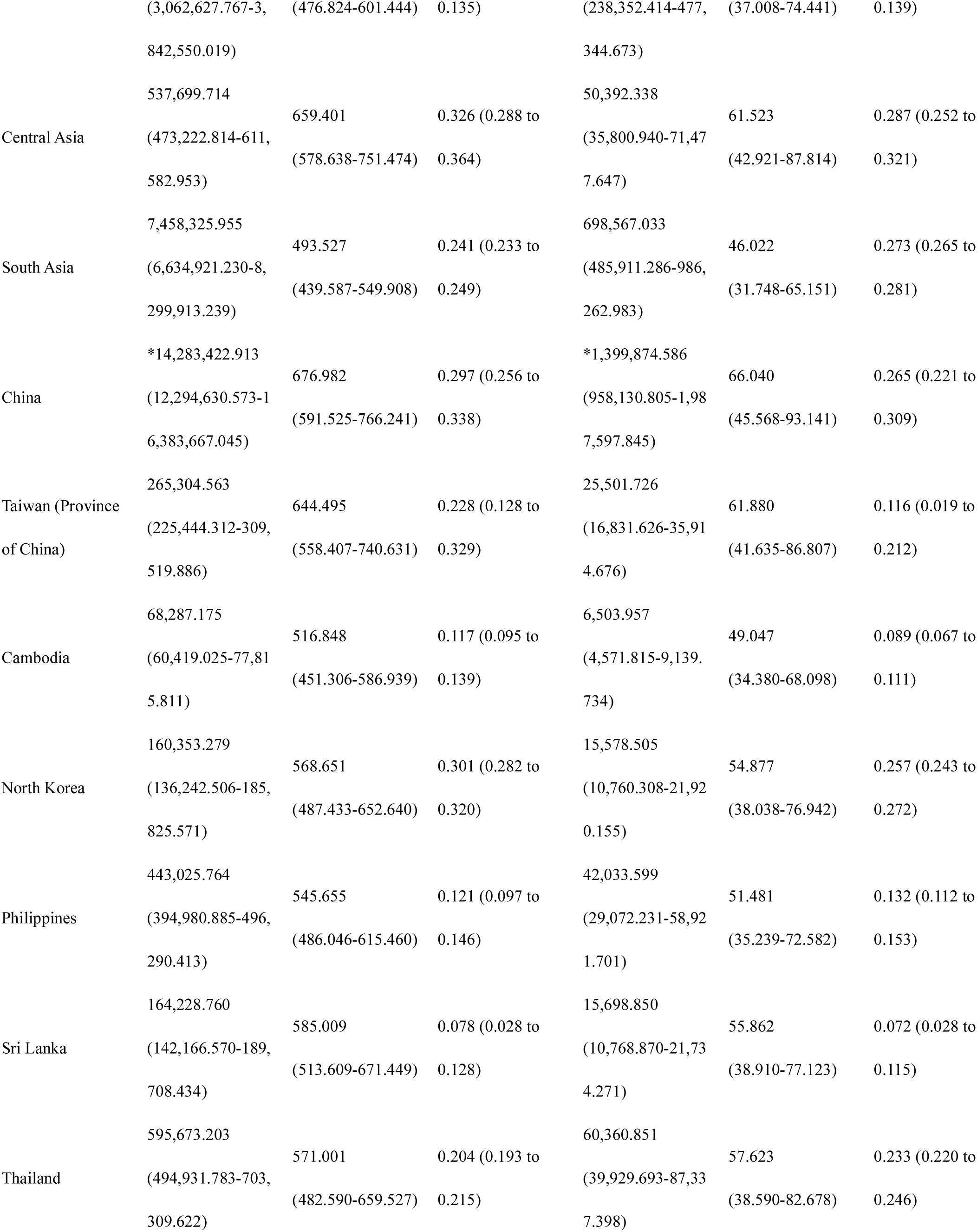
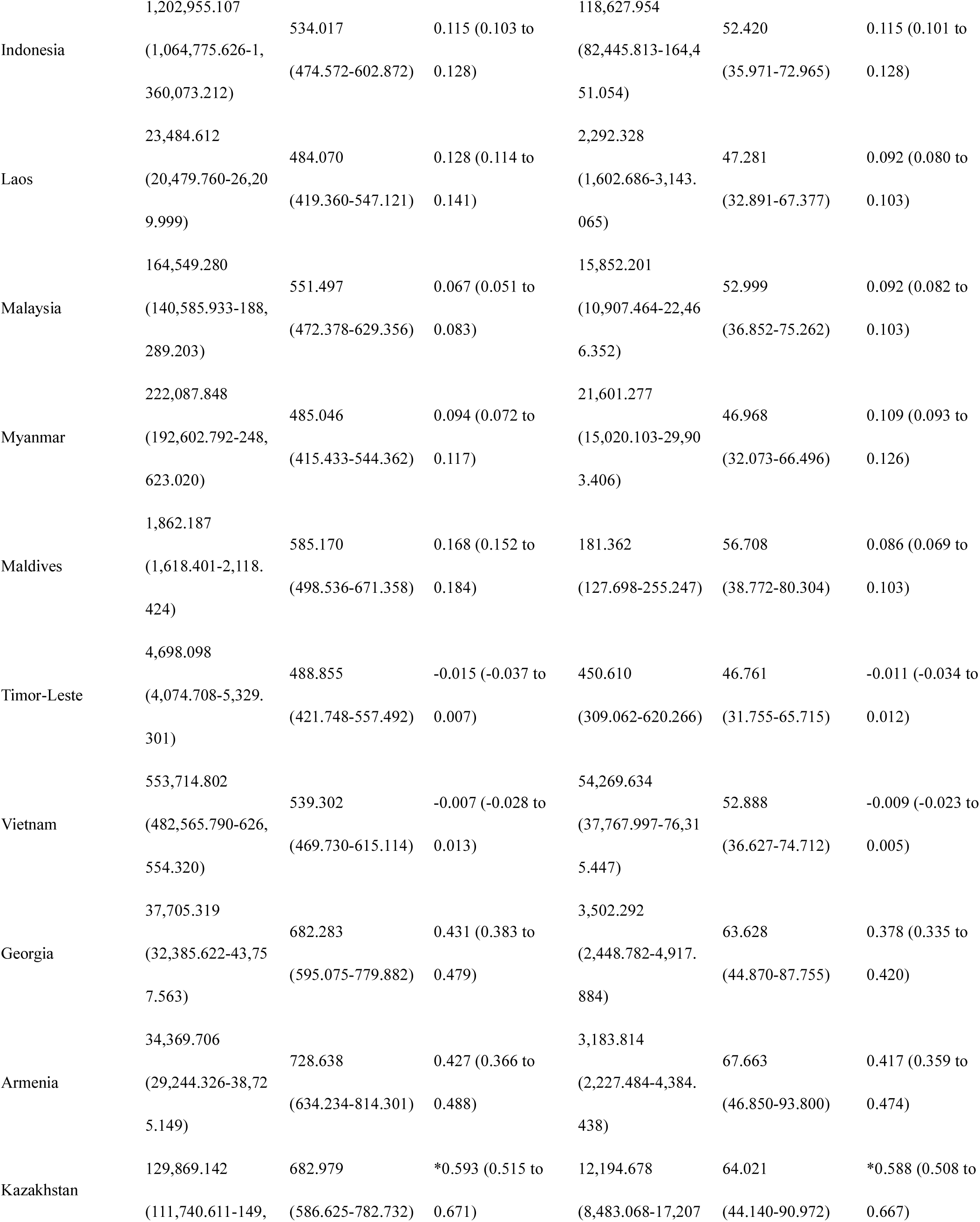
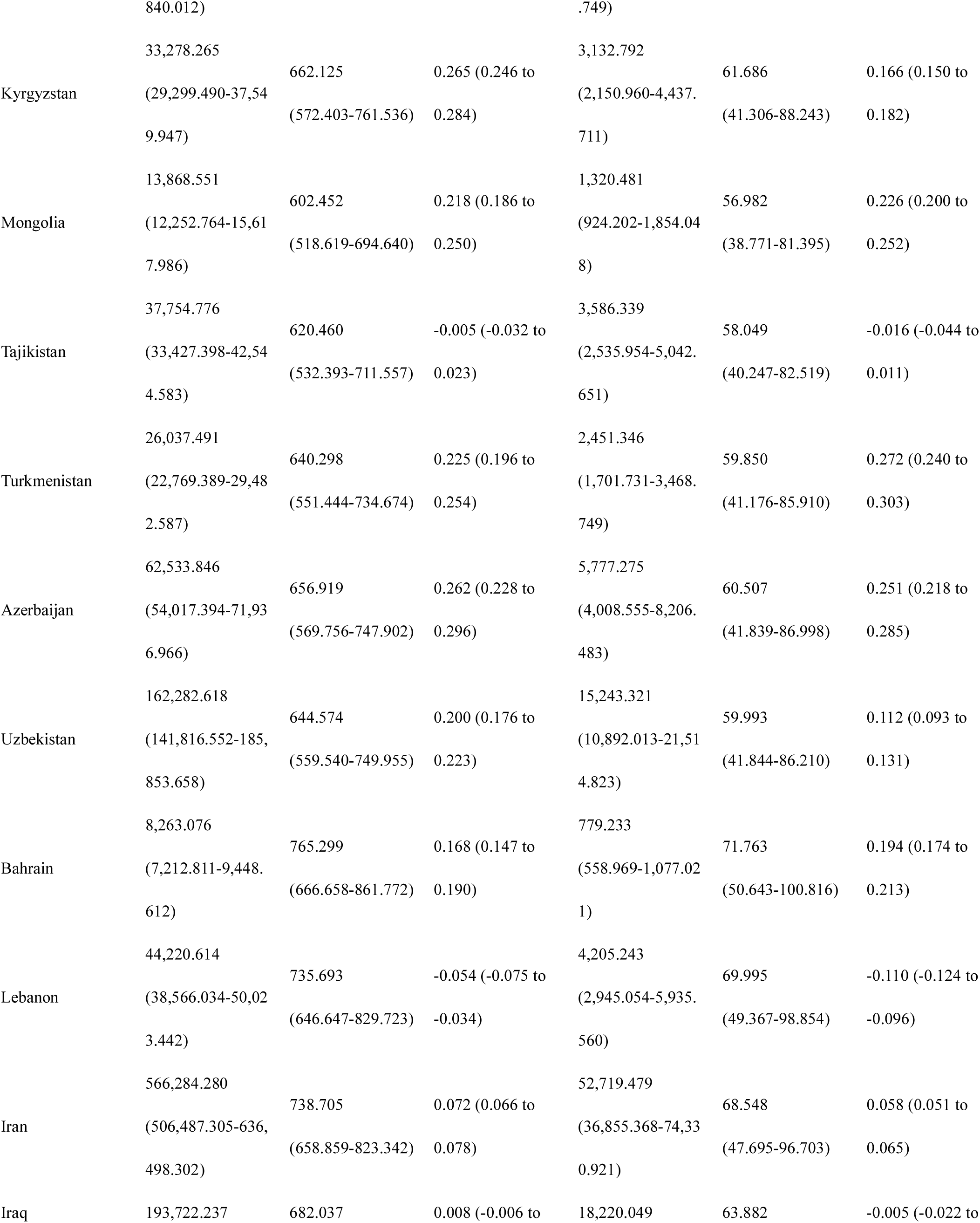
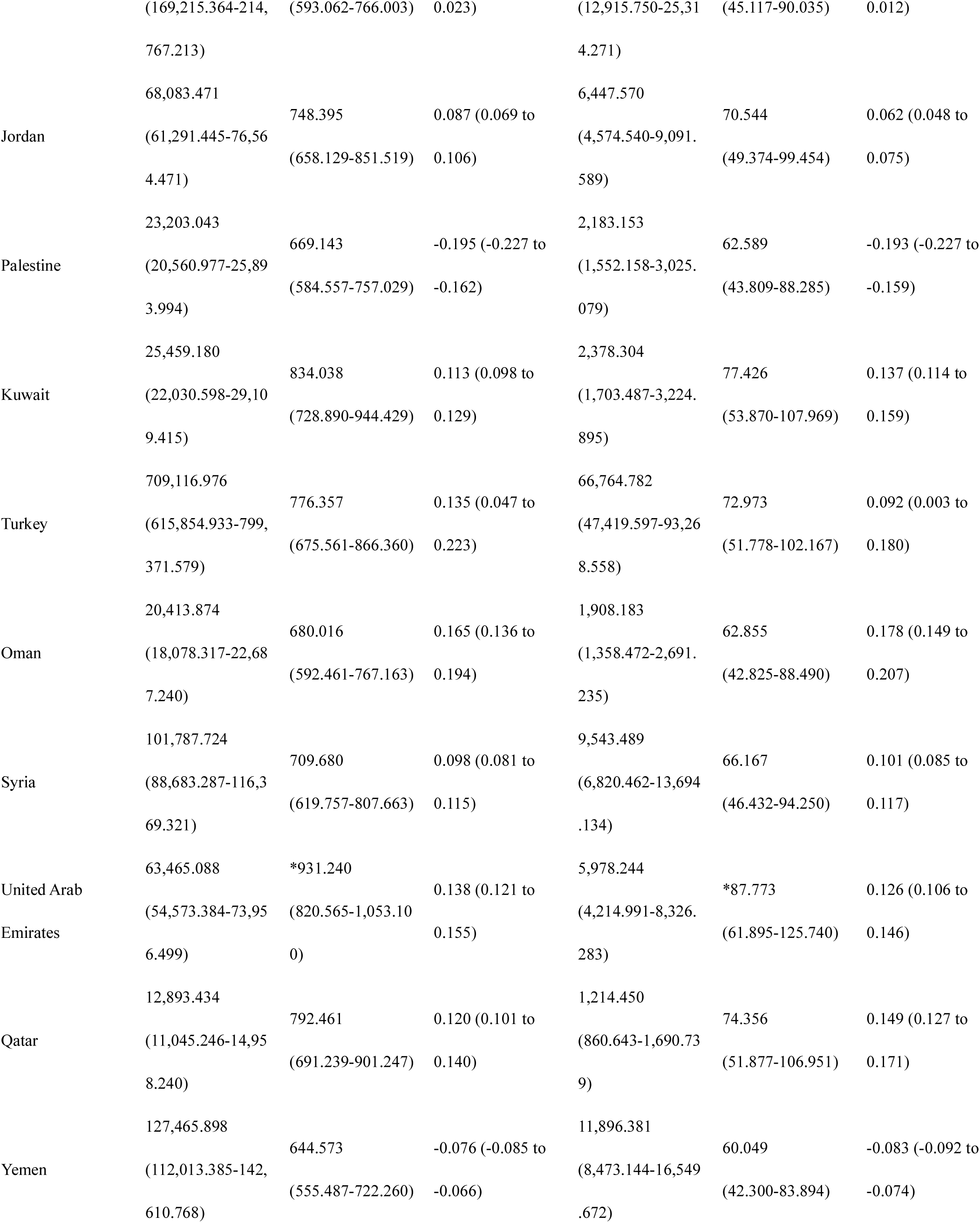
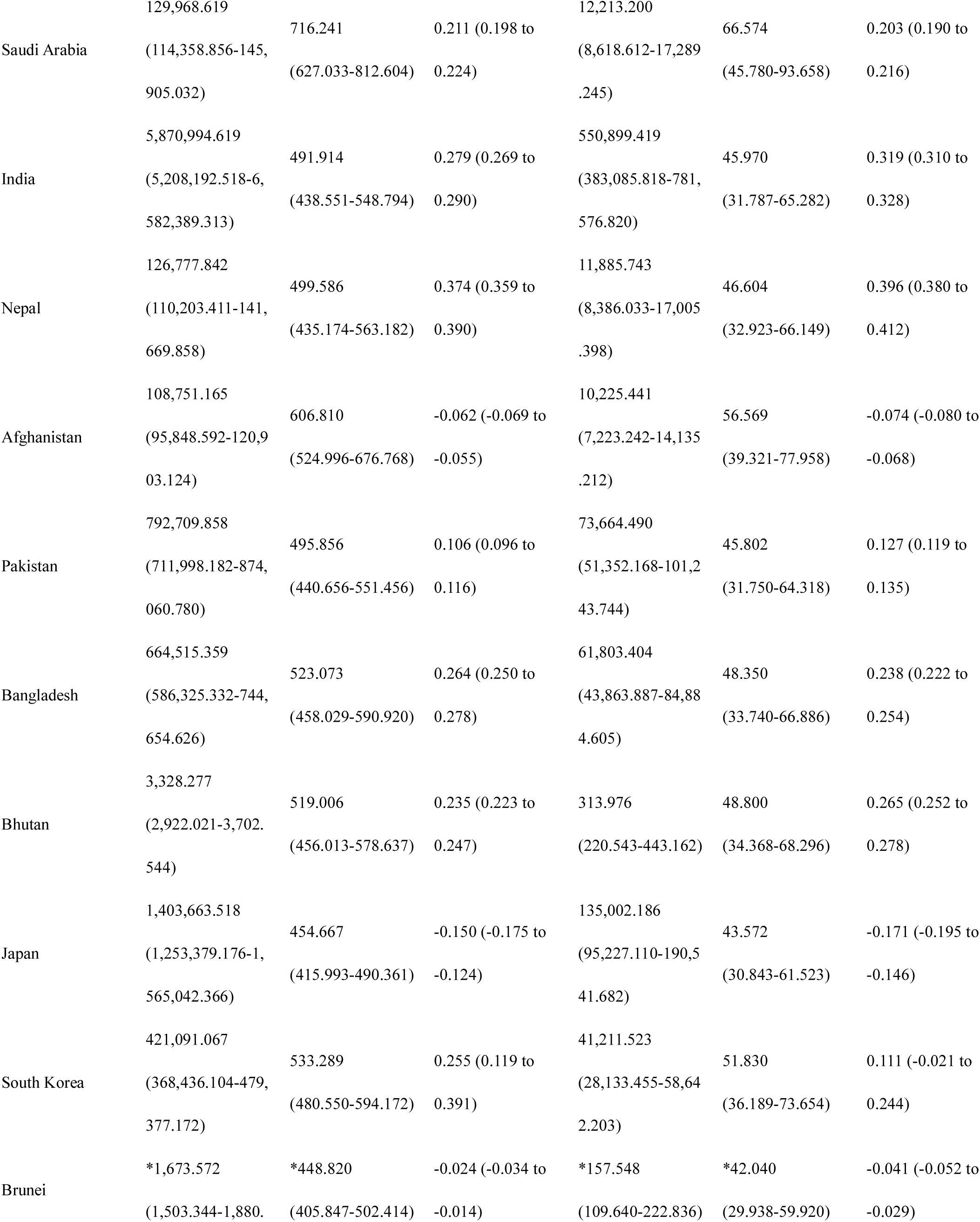
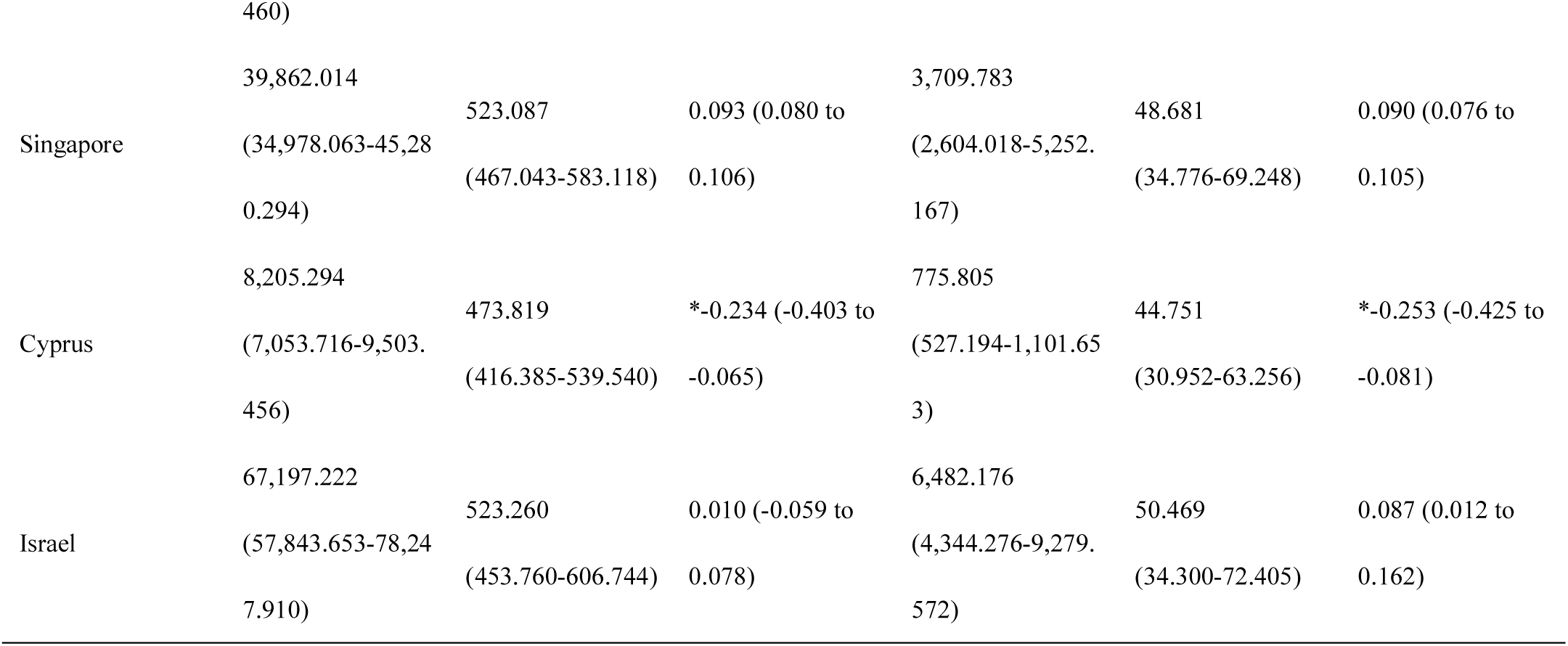
Heart failure prevalence cases, Years Lived with Disability (YLDs), age-standardized prevalence rates, YLDs rates, and estimated annual percentage changes from 1990-2023 for Asia, four Asian subregions, and 49 Asian countries (territories) in 2023. The four subregions in Asia are East Asia, Central Asia, South Asia, and Southeast Asia.

Disease burden change trends from 1990-2023 showed significant regional differences (Figure 1A-B). Heart failure ASPR in Asia showed an overall upward trend (Estimated Annual Percent Change [EAPC] = 0.252%), with Kazakhstan showing the fastest growth in age-standardized prevalence rates (EAPC = 0.593%), followed by Georgia (0.431%) and Armenia (0.427%), while Cyprus (-0.234%), Palestine (-0.195%), and Japan (-0.150%) showed declining trends. Age-standardized YLDs rates for heart failure in Asia showed an upward trend (EAPC = 0.238%), with Kazakhstan (0.588%), Armenia (0.417%), and Nepal (0.396%) showing the most significant growth, while Cyprus (-0.253%), Palestine (-0.193%), and Japan (-0.171%) showed declining trends.

**Figure 1.**
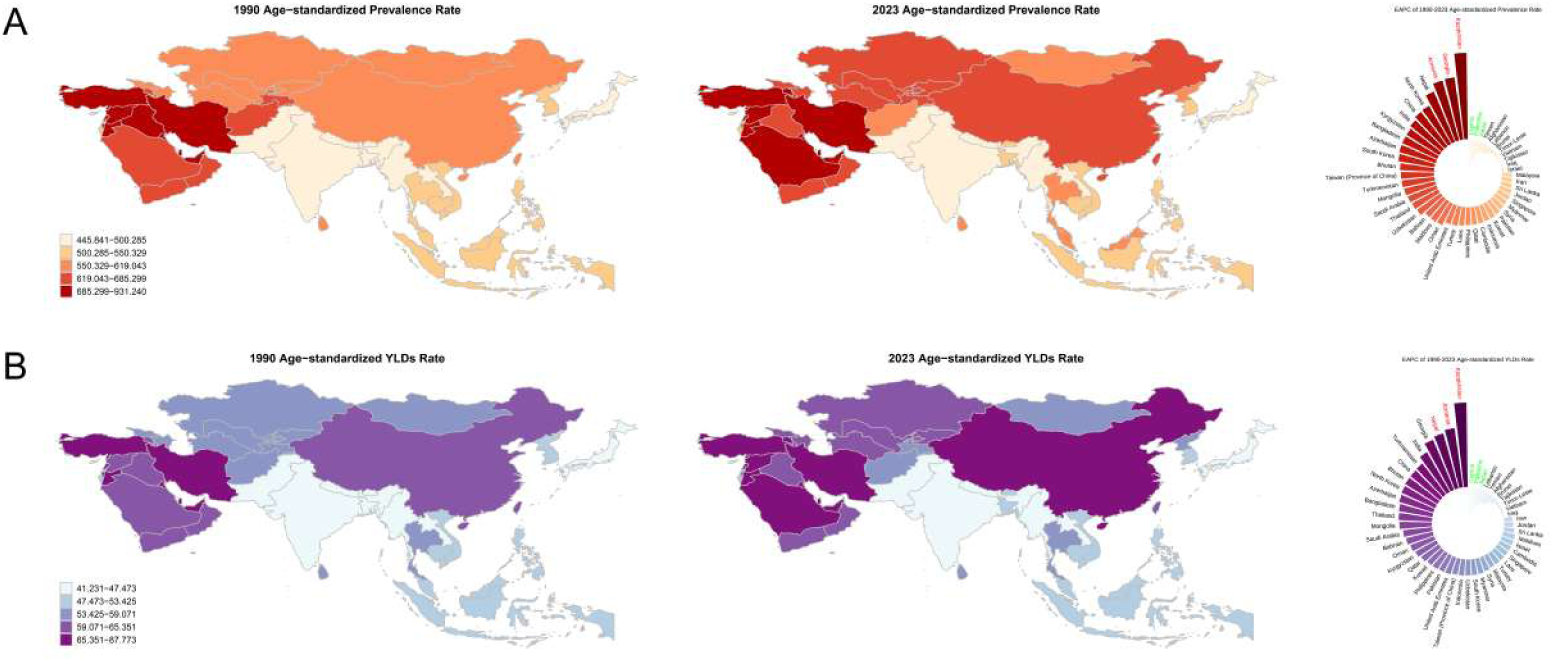
Trends of heart failure disease burden in Asia (1990-2023). Distribution and changes in age-standardized (per 100,000 population) prevalence rates (A) and YLDs rates (B) of heart failure in Asia. Each indicator includes three subplots: left panel shows 1990 distribution, middle panel shows 2023 distribution, right panel shows estimated annual percentage change (EAPC) from 1990 to 2023. Color intensity represents disease burden levels. In EAPC plots, red indicates the three countries with fastest increasing disease burden, green indicates the three countries with fastest declining burden.

### 3.2 Age and Gender Distribution Characteristics of Heart Failure Disease Burden

Age-gender specific analysis results showed that heart failure disease burden in Asia exhibited distinct age and gender distribution characteristics. In terms of absolute numbers, prevalence cases peaked in elderly age groups, with males reaching maximum values of approximately 2,567,585 cases in the 70-74 age group and females peaking at approximately 2,279,960 cases in the 70-74 age group (Figure 2A). The prevalence rates of heart failure increased significantly with age and male prevalence rates were higher than female prevalence rates. Prevalence rates reached highest levels in the 95+ age group, with males at 12,290.816/100,000 and females at 12,135.483/100,000 (Figure 2B).

**Figure 2.**
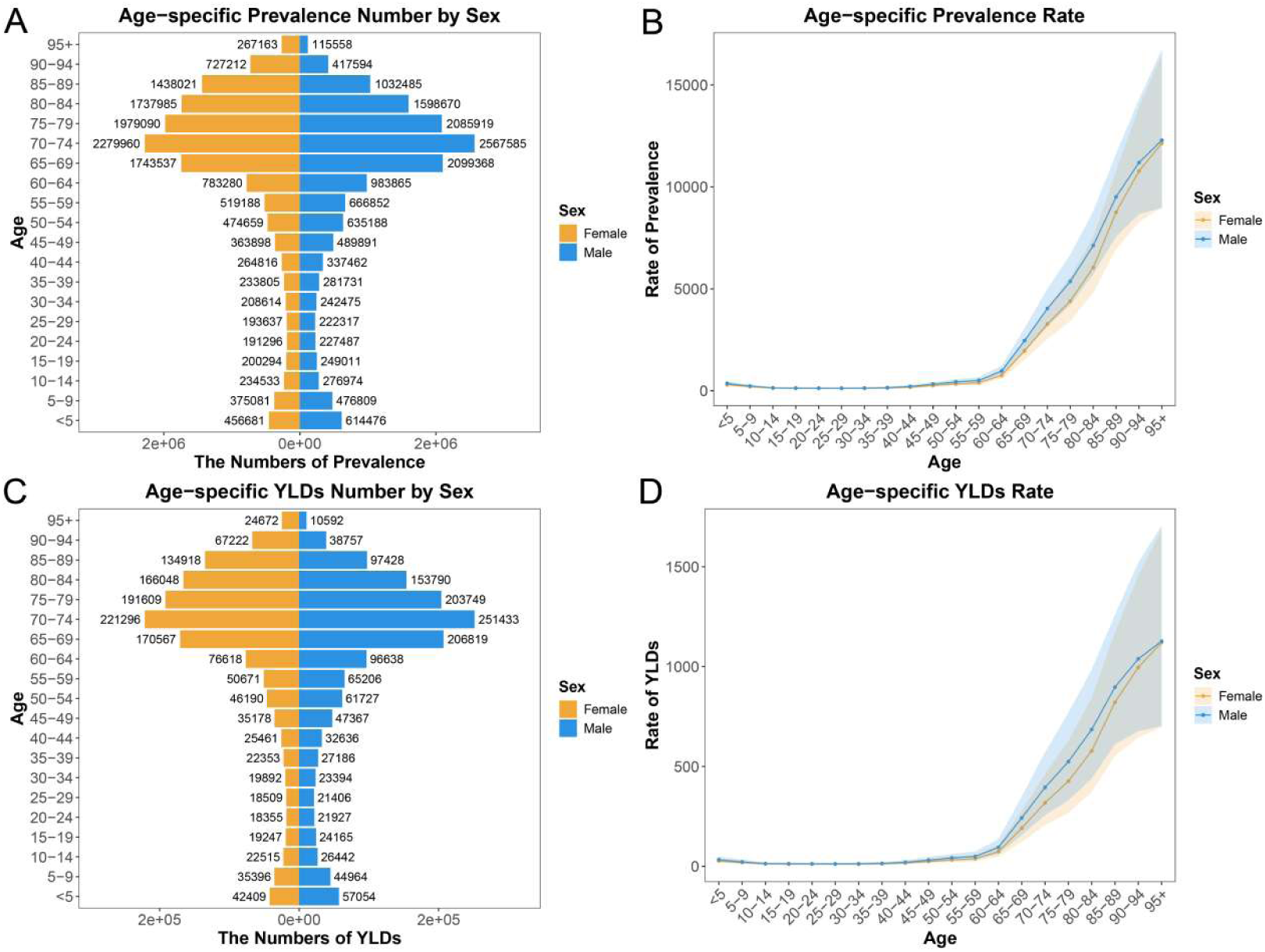
Age-gender specific distribution characteristics of heart failure disease burden in Asia. (A-B) Age-gender specific heart failure prevalence cases and prevalence rates. (C-D) Age-gender specific YLDs numbers and YLDs rates caused by heart failure.

YLDs absolute numbers showed males peaking at 251,433 in the 70-74 age group and females peaking at 221,296 in the 70-74 age group (Figure 2C). YLDs rates increased with age, reaching peaks in the 95+ age group with males at 1,126.582/100,000 and females at 1,120.669/100,000, with male prevalence rates higher than female rates (Figure 2D).

### 3.3 Joinpoint Regression Analysis

Joinpoint regression analysis results showed that both ASPR and age-standardized YLDs rates of heart failure in Asia from 1990-2023 exhibited overall upward characteristics, but the four subregions in Asia showed differentiated epidemic trends. Overall ASPR in Asia began slowly rising in 1990-1995 (APC = 0.192%), rapidly increased in 1996-1998 (APC = 0.650%), briefly declined in 1999-2004 (APC = -0.146%), accelerated upward in 2005-2014 (APC = 0.488%), and reached significant upward speed in 2019-2023 (APC = 0.302%). Male ASPR consistently exceeded female rates, with most significant increases in 1995-1999 (APC = 0.718%), while females showed sustained upward trends in 2013-2023 (APC = 0.437%) (Figure 3A).

**Figure 3.**
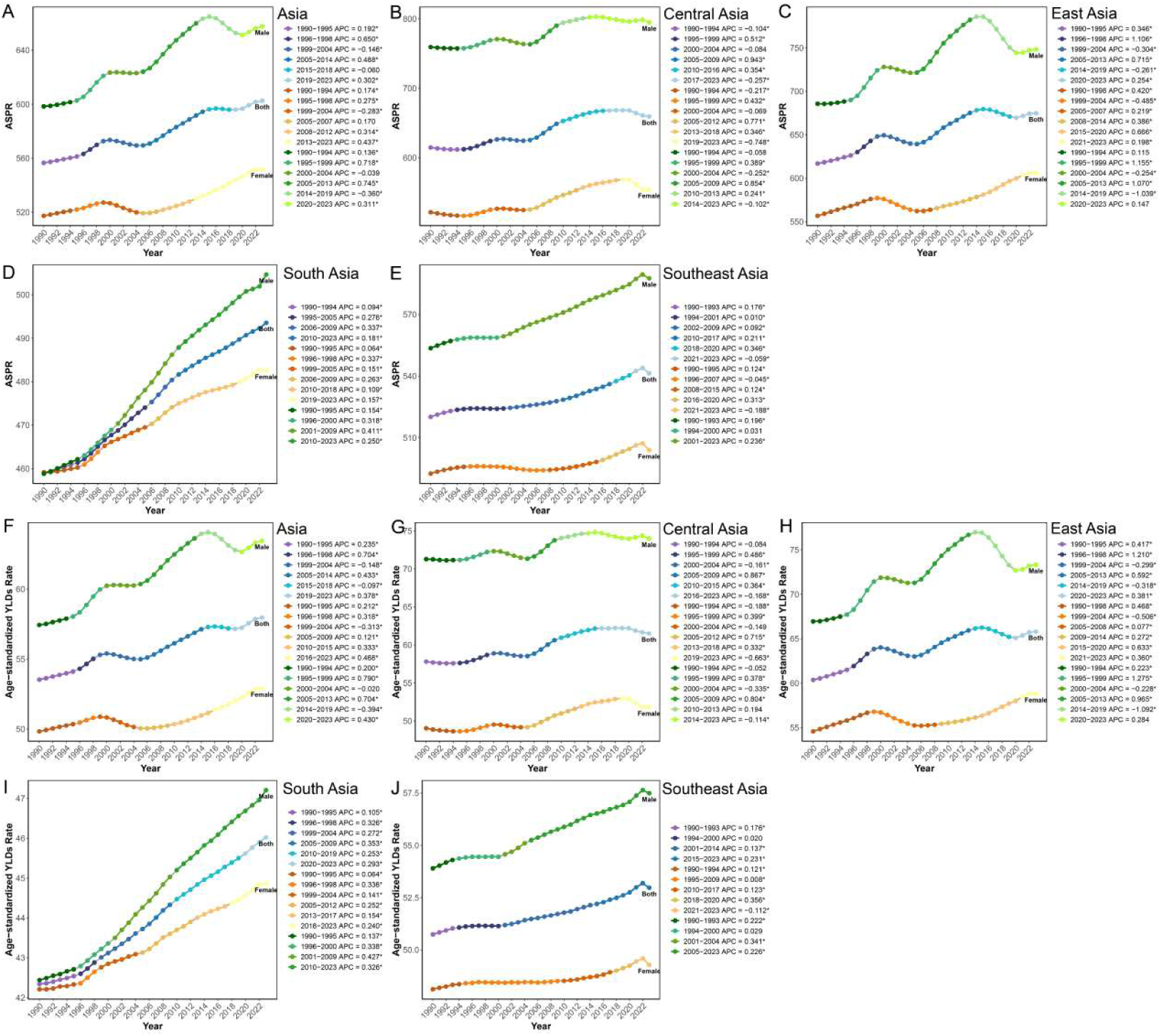
Joinpoint regression analysis of heart failure ASPR (A-E) and age-standardized YLDs rates (F-J) in Asia and four subregions from 1990-2023. The four Asian subregions are East Asia, Central Asia, South Asia, and Southeast Asia.

Age-standardized YLDs rate changes showed similar upward patterns, with overall Asia beginning to rise in 1990-1995 (APC = 0.235%), rapidly growing in 1996-1998 (APC = 0.704%), briefly declining in 1999-2004 (APC = -0.148%), significantly rising in 2005-2014 (APC = 0.433%), and showing upward trends in 2019-2023 (APC = 0.378%). Males showed most significant increases in 1995-1999 (APC = 0.790%), while females showed larger increases in 2016-2023 (APC = 0.468%) (Figure 3B).

### 3.4 Age-Period-Cohort Effect Analysis of Heart Failure

Age-Period-Cohort analysis revealed multidimensional changes in heart failure epidemic characteristics (Figure 4A-B). Age effects showed all indicators significantly increased with age, with prevalence rates continuously rising from 218.974/100,000 in the 2.5-year group to 16,612.715/100,000 in the 97.5-year group; YLDs rates continuously increased from 20.481/100,000 in the 2.5-year group to 1,509.201/100,000 in the 97.5-year group (Figure 4A-B). Period effect analysis showed that, using 2006.5 as reference (period rate ratio [PRR] = 1), prevalence risk and YLDs risk showed overall upward trends during the study period, with prevalence risk rising from 0.967 in 1996.5 to 1.083 in 2021.5. YLDs risk rose from 0.965 in 1996.5 to 1.078 in 2021.5.

**Figure 4.**
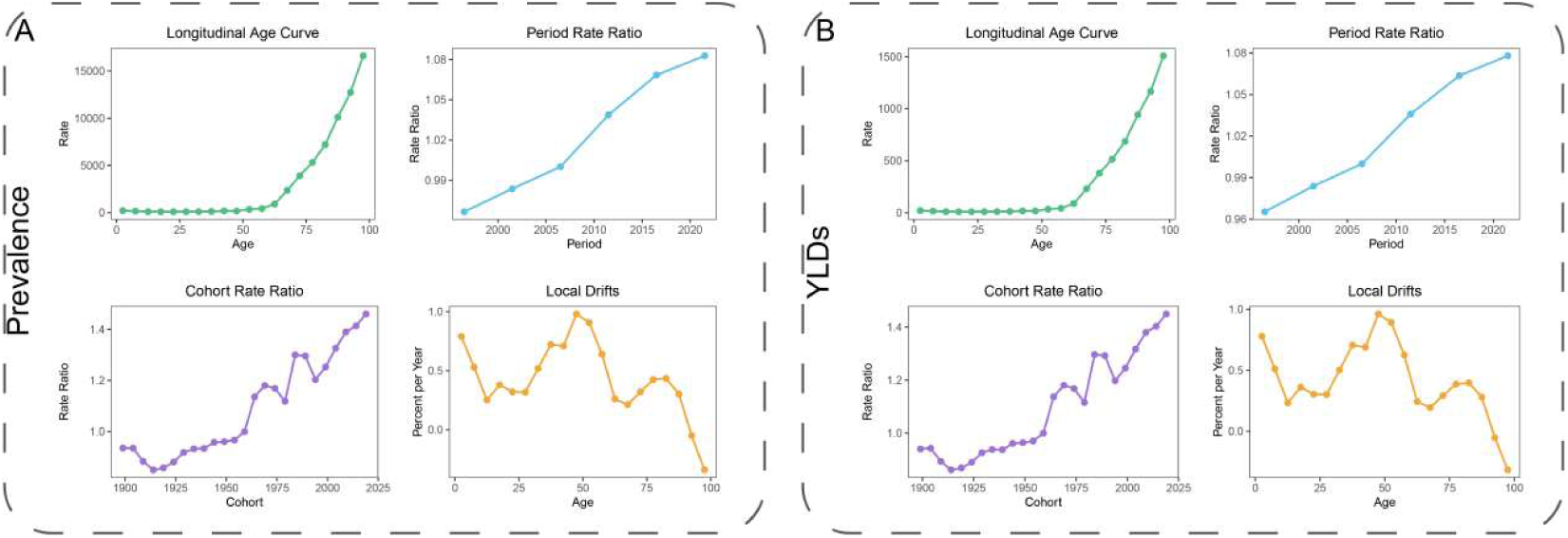
Age-Period-Cohort effect analysis of heart failure disease in Asia (1990-2023). (A-B) show Age-Period-Cohort effects for heart failure prevalence and YLDs, respectively. Upper left: Age effect plot showing relative risk changes of each indicator with age. Upper right: Period effect plot with 2006.5 as reference, showing relative risks at different time points. Lower left: Cohort effect plot with 1959 birth cohort as reference, reflecting cumulative effects of long-term factors such as lifestyle and environmental exposure. Lower right: Local drift plot showing annual percentage changes by age group, revealing dynamic change trends in disease burden across different age populations.

Cohort effect analysis indicated that, using the 1959 birth cohort as reference (cohort rate ratio [CRR] = 1), early birth cohorts (such as 1899) had relatively lower prevalence and YLDs risks (CRR = 0.936 and 0.940), while recent birth cohorts showed significantly increased risks (2019 prevalence CRR = 1.460, YLDs CRR = 1.450), reflecting overall trends of continuously rising heart failure risk across generations.

Local drift analysis showed prevalence risk exhibited positive growth in most age groups (0.212% to 0.981%), with larger increases in middle-age groups, while only the 92.5 and 97.5-year groups showed negative growth (-0.050% and -0.341%). YLDs risk increased in the 2.5-87.5 groups, with maximum increase in the 47.5-year group (0.962%), followed by decreased YLDs risk in the 92.5-97.5-year groups.

### 3.5 Decomposition Analysis of Heart Failure Disease Burden

Das Gupta decomposition analysis revealed that heart failure prevalence burden in Asia showed upward trends from 1990-2023, primarily driven by aging (49.04%), followed by population growth (41.80%), with epidemiological effects contributing positively (9.16%). Gender differences in driving factor composition were slight, with male prevalence burden changes dominated by aging (47.02%) followed by population growth (42.15%), while females similarly showed aging dominance (51.26%) followed by population growth (41.48%). Significant differences existed across the four subregions in Asia, with East Asia showing the largest aging contribution (73.75%), while Central Asia and South Asia were dominated by population growth (59.72% and 62.27%), and positive contributions from epidemiological effects increased from 8.18% in East Asia to 11.64% in Central Asia, reflecting differences in disease epidemic characteristic changes across regions with different development levels (Figure 5A-B).

**Figure 5.**
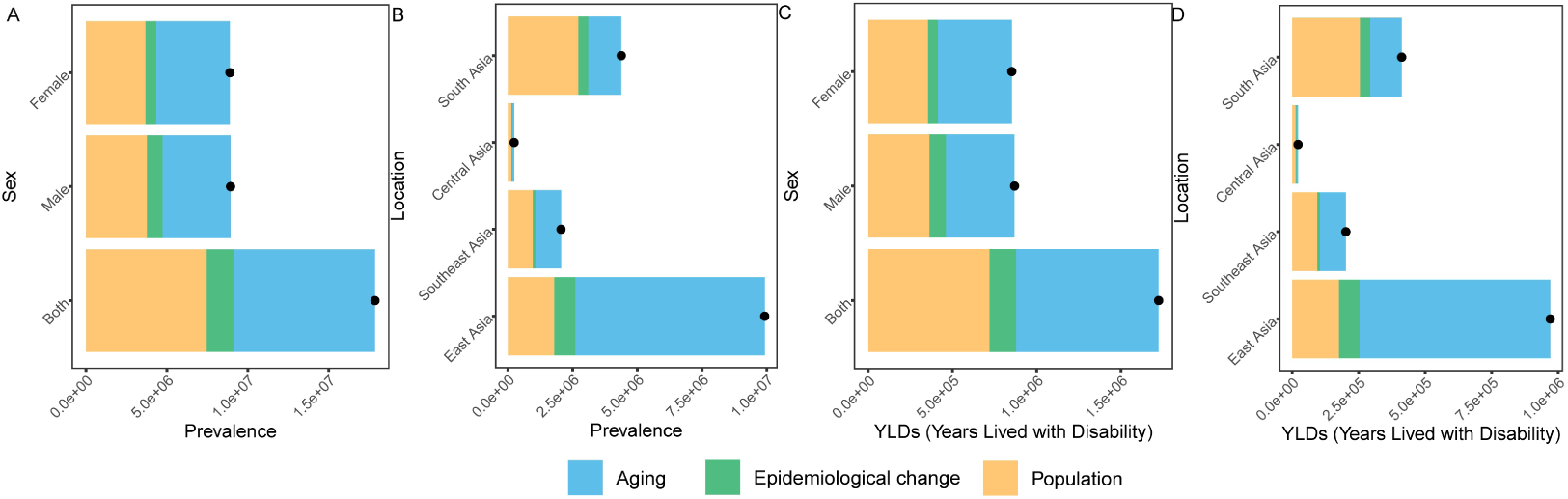
Decomposition analysis of heart failure prevalence (A, B) and YLDs (C, D) burden changes in Asia and four subregions from 1990-2023. The four Asian subregions are East Asia, Central Asia, South Asia, and Southeast Asia.

YLDs burden similarly showed upward trends, with Asia-level changes primarily driven by aging (49.14%), followed by population growth (41.75%) and epidemiological effects (9.11%). Gender differences were slight, with aging contributing slightly more to females (51.55% vs. 46.97%), while epidemiological effects showed more prominent positive contributions to males (11.15% vs. 6.79%). Four subregional in Asia stratification showed East Asia YLDs burden increases were primarily driven by aging (73.95%), with positive epidemiological effect contributions (7.92%). Central Asia and South Asia YLDs burden increases were primarily driven by population growth (60.89% and 61.88%) (Figure 5C-D).

### 3.6 Frontier Analysis

Frontier analysis results indicated significant nonlinear relationships between heart failure age-standardized YLDs rates and Human Development Index (HDI) (Figure 6A). Specifically, age-standardized YLDs rates in most countries showed slight increases with HDI increases, and most countries deviated from frontier theoretical optimal values, indicating substantial room for improvement in heart failure control in these countries.

**Figure 6.**
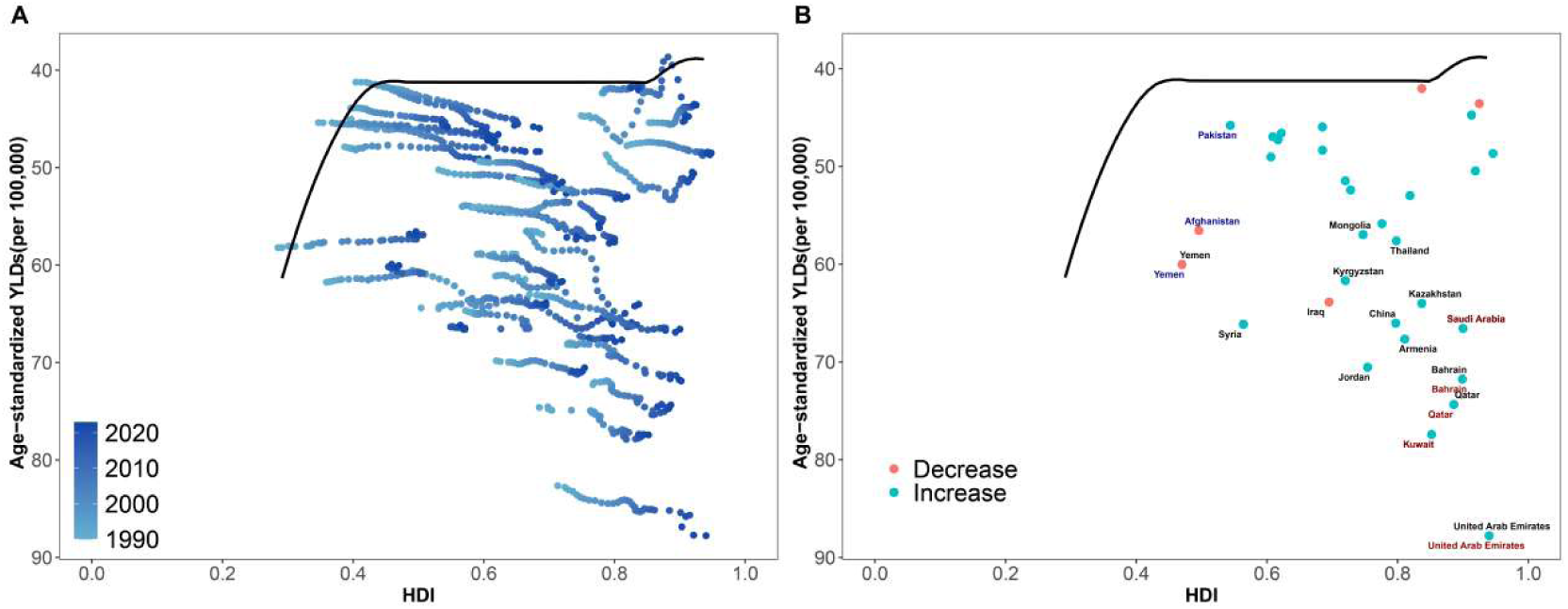
Frontier analysis of heart failure disease burden in 32 countries from 1990-2023 based on HDI and age-standardized YLDs rates. (A) Scatter plot of HDI versus heart failure age-standardized YLDs rates for 32 countries from 1990-2023. Black curve represents frontier production function fitting curve, indicating minimum YLDs rates at given HDI levels. Different colored points represent different years. (B) Change trends in heart failure age-standardized YLDs rates for selected countries in 2023. Black solid line represents frontier production function fitting curve, colored dotted lines represent YLDs rate changes for individual countries. Black text indicates the 15 points with largest distance differences; blue text indicates the 5 countries with smallest distance differences among low HDI countries; red text indicates the 5 countries with largest distance differences among high HDI countries.

Cross-sectional analysis results for 2023 showed that actual age-standardized YLDs rates in multiple countries significantly deviated from DEA efficiency frontiers corresponding to their HDI levels, indicating substantial untapped potential in disease prevention and control under existing social development conditions (Figure 6B). High HDI countries including United Arab Emirates, Kuwait, Qatar, and Bahrain showed poor control of age-standardized YLDs rates. Countries including Syria, Mongolia, Kazakhstan, and China showed particularly significant deviations between actual age-standardized YLDs rates and theoretical optimal values.

### 3.7 Health Inequality Analysis

Health inequality analysis results showed that health inequality patterns for heart failure from 1990 to 2023 exhibited significant deterioration trends. Slope Index of Inequality (SII) showed sustained deterioration, rising dramatically from 10.684 in 1990 to 25.003 in 2023, with deterioration amplitude reaching 134.0%. SII values showed clear upward trajectories throughout the observation period, with particularly significant deterioration after 2010, rapidly climbing from 13.811, indicating continuously expanding absolute differences in heart failure disease burden across regions with different HDI levels (Figure 7A-B).

**Figure 7.**
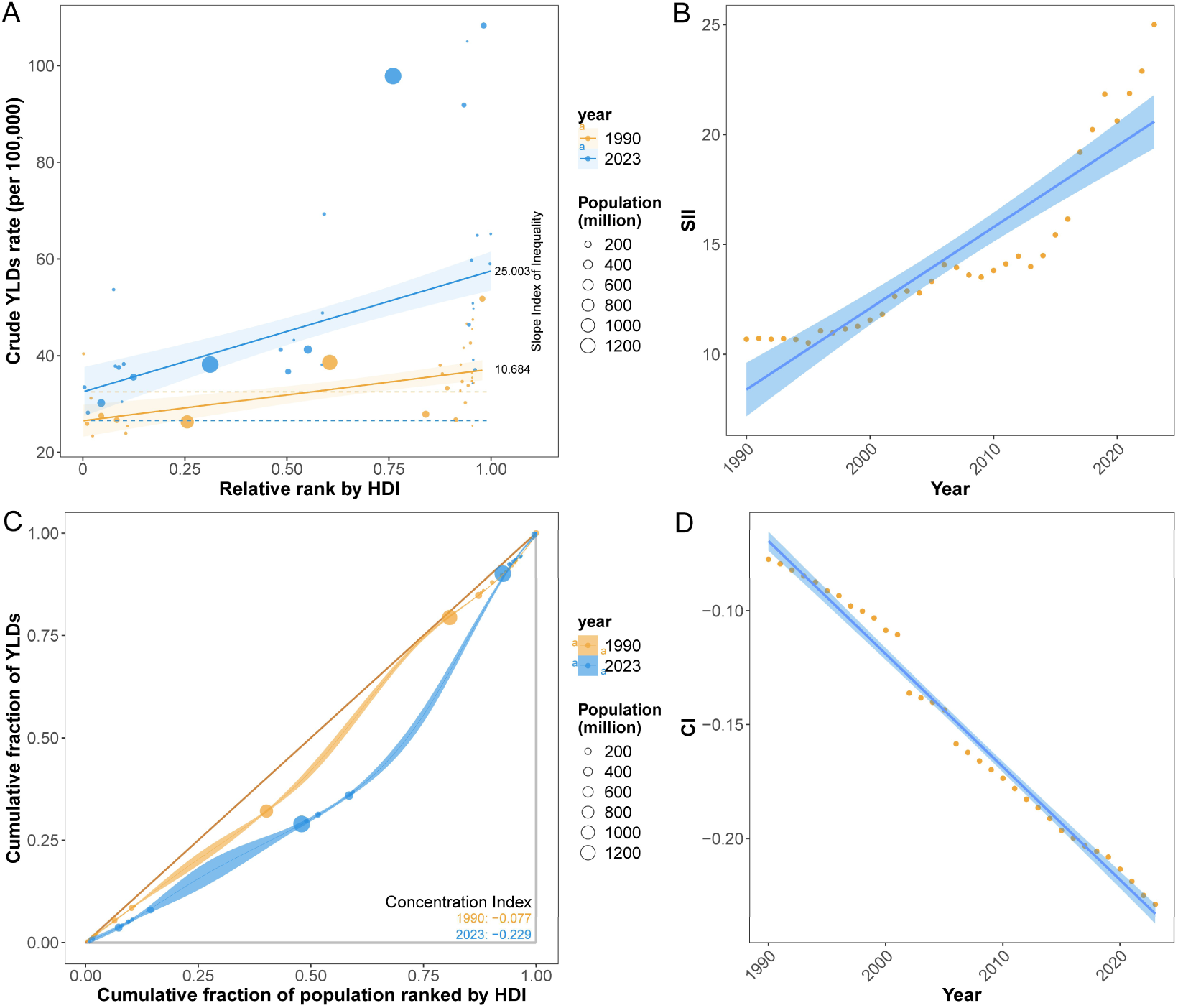
Health inequality analysis of heart failure YLDs rates from 1990-2023. (A) Trend plot of Slope Index changes for heart failure YLDs rates from 1990-2023. The value of SII was 10.684 in 1990 and 25.003 in 2023. (B) Scatter plot of Slope Index for heart failure YLDs rates from 1990-2023. (C) Inequality curves for heart failure YLDs rates. The value of CI was -0.077 in 1990 and -0.229 in 2023. (D) Scatter plot of CI for heart failure YLDs rates from 1990-2023.

Concentration Index (CI) maintained negative value characteristics throughout the study period, gradually declining from -0.077 in 1990 to -0.229 in 2023. CI values showed sustained deterioration patterns, slowly declining from 1990-2023 with continuously increasing absolute values, indicating heart failure disease burden was disproportionately concentrated in low HDI regions (Figure 7C-D).

### 3.8 Heart Failure Disease Burden Predictions Based on BAPC Models

Based on the continuation of historical trends, we conducted a BAPC model prediction. Prediction results showed that ASPR in Asia is projected to rise from 602.601/100,000 in 2023 to 623.351/100,000 in 2038. The four subregions in Asia showed significant differences: East Asia is projected to rise from 680.934/100,000 in 2023 to 691.842/100,000 in 2038; South Asia is expected to rise from 496.971/100,000 in 2023 to 516.154/100,000 in 2038; Central Asia is projected to show declining trends, from 664.334/100,000 in 2023 to 637.481/100,000 in 2038; Southeast Asia similarly shows slight declines, from 545.631/100,000 in 2023 to 537.901/100,000 in 2038 (Figure 8A-E).

**Figure 8.**
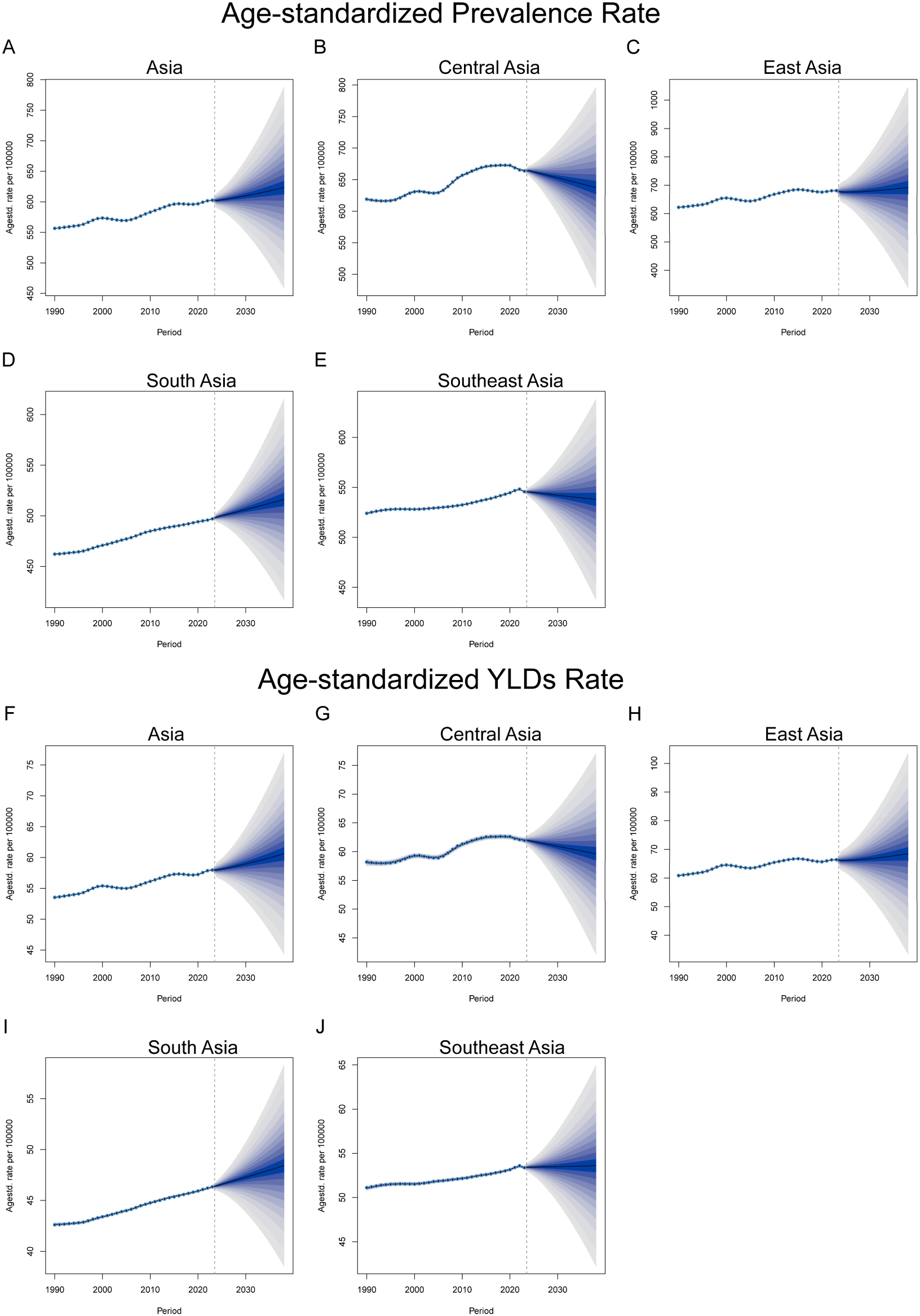
BAPC model-based predictions for 2024-2038 heart failure age-standardized prevalence (A-E) and YLDs (F-J) in Asia and four subregions. The four Asian subregions are East Asia, Central Asia, South Asia, and Southeast Asia.

Age-standardized YLDs rate predictions showed Asia will rise from 57.968/100,000 in 2023 to 60.596/100,000 in 2038. The four subregions in Asia change patterns were similar to prevalence rates but with different magnitudes: East Asia from 66.384/100,000 in 2023 to 68.435/100,000 in 2038; South Asia from 46.316/100,000 in 2023 to 48.411/100,000 in 2038; Central Asia showing declining trends from 61.996/100,000 in 2023 to 59.613/100,000 in 2038; Southeast Asia showing minimal changes from 53.398/100,000 in 2023 to 53.596/100,000 in 2038 (Figure 8F-J).

## 4 Discussion

This study found that the ASPR and YLDs rate of heart failure in Asia demonstrated a continuous upward trend from 1990 to 2023, with projections indicating that this trend will persist over the next 15 years. This finding is largely consistent with global heart failure epidemiological studies ^[5, 11]^, reflecting the increasingly severe public health challenge posed by cardiovascular diseases in Asia.

The geographical disparity analysis revealed significant spatial heterogeneity in the heart failure disease burden. East Asia exhibited the highest ASPR (674.8/100,000) and YLDs rate (65.8/100,000) in Asia, while South Asia showed relatively lower rates. This geographical distribution pattern may be attributed to multiple factors. First, the rapid economic development and urbanization in East Asia have led to the westernization of lifestyles, with factors such as high-salt diets, sedentary lifestyles, and increased work-related stress promoting the prevalence of heart failure risk factors including hypertension and diabetes ^[12–14]^. Second, East Asia is experiencing a more rapid population aging process. Decomposition analysis revealed that 73.75% of the disease burden increase in East Asia was attributable to aging, substantially higher than in Central and South Asia, which is consistent with previous reports indicating that East Asia has one of the fastest aging rates globally ^[15]^. Third, variations in the development level of healthcare systems and disease diagnostic capabilities across different regions may also influence the reporting rates of disease burden. The relatively well-established healthcare systems in East Asia may have improved the diagnosis and survival rates of heart failure, thereby increasing the statistical values of prevalence and YLDs ^[16, 17]^. Notably, high-income countries such as the United Arab Emirates exhibited exceptionally high disease burdens, which may be closely associated with rapid economic transformation, high urbanization levels, high prevalence of obesity and diabetes, and the shift from traditional dietary patterns to high-calorie Western diets ^[18, 19]^.

It is essential to consider the potential influence of contemporary heart failure pharmacotherapies on the observed trends. Over the past decade, landmark trials have established the clinical benefits of angiotensin receptor-neprilysin inhibitors (ARNi, e.g., sacubitril/valsartan) and sodium-glucose cotransporter-2 (SGLT2) inhibitors (e.g., dapagliflozin, empagliflozin) in reducing hospitalization and mortality in patients with heart failure ^[20–22]^. The widespread adoption of these agents, particularly in high-income Asian countries such as Japan, South Korea, and Singapore, may have contributed to improved survival and consequently increased prevalence by extending the duration of disease. However, the uptake of these therapies remains highly uneven across Asia, with limited access in low- and middle-income countries due to cost, regulatory, and healthcare infrastructure barriers ^[23]^. The GBD modeling framework does not explicitly incorporate treatment-specific parameters, meaning that the effects of therapeutic advances are captured only indirectly through their aggregate impact on cause-specific mortality and disability weights. This limitation should be considered when interpreting the observed trends, as accelerating adoption of guideline-directed medical therapy could potentially attenuate the projected increases in heart failure burden.

Gender disparity analysis demonstrated that the heart failure disease burden in males consistently exceeded that in females, a finding concordant with multiple global epidemiological studies ^[24, 25]^. The higher disease burden in males may be related to various factors. At the biological level, the protective effects of estrogen on the cardiovascular system play an important role in premenopausal women, including mechanisms such as improving endothelial function and reducing oxidative stress and inflammatory responses ^[26–28]^. At the social-behavioral level, smoking rates and alcohol consumption are generally higher among males, with more prevalent occupational stress exposure and unhealthy lifestyles, which collectively increase the risk of developing heart failure precursor conditions such as ischemic heart disease and hypertension ^[29]^. However, Joinpoint regression analysis revealed that the annual percentage change (APC) in ASPR for females (0.437%) exceeded that of males from 2013 to 2023, suggesting an accelerating heart failure burden among women. This may be associated with rapid lifestyle changes among Asian women, rising smoking rates, and diminished protective effects post-menopause ^[30]^.

Health inequality analysis revealed that the slope index of inequality deteriorated by 134% from 1990 to 2023, with the negative concentration index continuously expanding, indicating that the heart failure disease burden is increasingly and disproportionately concentrated in low-HDI regions. This widening health inequality reflects the exacerbation of developmental imbalances within Asia. Low-income regions face a dual burden when addressing the epidemiological transition of cardiovascular diseases—confronting both infectious disease challenges and the rapid growth of non-communicable diseases—while insufficient medical resources and prevention systems limit their capacity for effective response ^[31, 32]^. Frontier analysis further demonstrated substantial variations in heart failure control efficiency among countries even at comparable HDI levels, suggesting that many countries still have significant room for improvement through optimized resource allocation and adoption of best practices.

The COVID-19 pandemic, which profoundly affected healthcare systems across Asia from 2020 to 2023, may have influenced heart failure burden estimates in several ways. First, disruption of routine healthcare services during pandemic waves likely led to delayed diagnosis and underreporting of heart failure, potentially causing underestimation of prevalence in the immediate pandemic period ^[33]^. Second, SARS-CoV-2 infection has been associated with direct myocardial injury, myocarditis, and the development of new-onset heart failure, which may have contributed to increased incident cases ^[34]^. Third, pandemic-related disruptions in medication adherence, rehabilitation services, and follow-up care may have worsened outcomes among existing heart failure patients, potentially increasing YLDs ^[35]^. Fourth, excess cardiovascular mortality during the pandemic may have paradoxically reduced prevalence by removing patients from the prevalent pool. These competing effects create complexity in interpreting trends during 2020–2023 and may also introduce bias into the BAPC projections, which extrapolate from data that include this atypical period. The GBD 2023 study incorporates COVID-19-related excess mortality estimates and adjusts for pandemic-related data disruptions, but residual distortion of trend estimates cannot be excluded.

Projections based on the BAPC model indicated that the ASPR of heart failure in Asia is expected to increase from 602.601/100,000 to 623.351/100,000, and the age-standardized YLDs rate from 57.968/100,000 to 60.596/100,000 from 2024 to 2038. This sustained upward trend is primarily driven by population aging and epidemiological transition, portending greater pressure on Asian healthcare systems from heart failure in the future ^[36]^. Notably, the four Asian sub-regions exhibited divergent trends: East and South Asia are projected to continue rising, while Central and Southeast Asia may experience slight declines. These differences may reflect varying progress in implementing cardiovascular disease prevention and control strategies and developing healthcare systems across regions ^[37, 38]^. This underscores the necessity for further interventions. Firstly, strengthening primary prevention of cardiovascular risk factors, particularly population-based interventions targeting modifiable risk factors such as hypertension, diabetes, and obesity. Secondly, improving early screening and hierarchical diagnosis and treatment systems for heart failure, and enhancing the diagnostic and management capabilities of primary healthcare institutions. Thirdly, emphasizing health equity by increasing resource investment in low-income regions and vulnerable populations to narrow health disparities ^[39]^. Finally, promoting cross-national cooperation and experience sharing, drawing on successful prevention and control experiences from other countries to enhance overall prevention and control efficiency.

The strength of this study lies in utilizing the latest GBD 2023 data and employing multiple advanced statistical methods to comprehensively analyze the multidimensional characteristics of heart failure disease burden in Asia. However, the study has certain limitations. First, although the GBD 2023 framework uses DisMod-MR 2.1 to enforce internal consistency and applies systematic bias corrections, the quality of input data varies substantially across Asian countries. Nations with less developed disease surveillance systems, vital registration infrastructure, or diagnostic capacity may contribute lower-quality data, resulting in wider uncertainty intervals and potentially less precise estimates. Underreporting or diagnostic misclassification of heart failure—particularly in low-income settings where echocardiographic confirmation is less available—may affect the accuracy of country-level prevalence and YLD estimates and should be considered when interpreting findings from these regions. Second, diagnostic criteria for heart failure may vary across countries and time periods, affecting data comparability. Third, the prediction model is based on extrapolation of historical trends and does not fully account for the potential effects of future policy interventions and advances in medical technology ^[40]^. The BAPC-based projections assume the continuation of historical trends in age, period, and cohort effects and cannot incorporate the potential impact of future events, including breakthroughs in heart failure therapeutics, implementation of new public health policies, shifts in risk factor prevalence due to behavioral or environmental changes, or unforeseen societal disruptions such as pandemics or economic crises. Consequently, these projections should be interpreted as indicative trend estimates rather than precise forecasts, and actual future burden may deviate substantially if the underlying determinants change.

## Conclusion

In conclusion, this study reveals the grave reality and future challenges of the heart failure disease burden in Asia, including widening health inequalities, significant geographical and gender disparities. Only through multi-level, multi-sectoral collaborative efforts can the rising trend of heart failure disease burden be effectively curbed and the overall cardiovascular health level in Asia be improved.

## Data Availability

GBD 2023

## Author Contributions

GGP contributed to the conceptualization and design of the study. GGP acquired and curated the GBD data. GGP and LL performed the formal analysis and visualization. GGP developed the methodology and conducted statistical modeling. GGP and YP wrote the original draft of the manuscript. All authors contributed to the review and editing of the manuscript. YP supervised the study and provided critical revisions. All authors have read and approved the final manuscript.

## Acknowledgments

We thank all the people who contributed to the data collection, analysis, vetting, and critical interpretation of the GBD study overall. Additionally, we would like to acknowledge the GBD region affiliates who provided the data used in this study.

## Data Availability Statement

The data used were publicly available for this study. The website of the data is https://vizhub.healthdata.org/gbd-results/.

